# Objective extraction of movement features prompting expert identification of dystonia in ambulatory children with cerebral palsy

**DOI:** 10.1101/2020.05.01.20086918

**Authors:** Bhooma R. Aravamuthan, Keisuke Ueda, Hanyang Miao, Laura Gilbert, Sarah Smith, Toni Pearson

**Affiliations:** Department of Neurology, Division of Pediatric Neurology, Washington University School of Medicine and St. Louis Children’s Hospital, St. Louis, Missouri, USA

## Abstract

**Aim:** To determine the specific movement features in children with cerebral palsy (CP) that prompt expert identification of dystonia

**Methods:** Dystonia identification in CP, particularly when co-morbid with spasticity, can be difficult. For this retrospective case-control study, we conducted a qualitative thematic analysis of consensus-building discussions between three pediatric movement disorders physicians as they attempted to identify the presence or absence of dystonia in gait videos of 40 subjects with spastic CP and periventricular leukomalacia.

**Results:** Initial unanimous consensus regarding the presence or absence of dystonia was achieved in 12 videos (30%). Following consensus building discussion, 22 additional videos (55%) yielded unanimous consensus. Two main themes were generated: 1) Unilateral leg or foot adduction that was variable over time, and 2) Difficulty in identifying dystonia. Codes contributing to Theme 1 were more likely to appear when a discussant was favoring the presence of dystonia in a video (Chi-square, p=0.004).

**Discussion:** These results provide specific movement features that could aid dystonia diagnosis in ambulatory children with CP. However, these results also suggest that, even amongst putative motor phenotyping experts, visual dystonia diagnosis in CP remains difficult, highlighting the need for developing and using objective dystonia diagnosis measures.

**What this paper adds:**

- Dystonia identification is visually difficult, even for experts
- Unilateral lower extremity variable adduction could represent gait dystonia in cerebral palsy
- Qualitative thematic analysis objectively identifies expert-cited dystonia features in cerebral palsy

## Introduction

Dystonia is characterized by intermittent or sustained postures that are triggered or worsened by voluntary movement or heightened arousal.^1^ It is the predominant form of abnormal tone in 15% of those with cerebral palsy (CP) and may be present to some degree in many more children with CP, particularly in the lower extremities.^2,3^ However, identification of dystonia in CP remains difficult.^4,5^ Perhaps due to its variable nature and common co-morbidity with spasticity in CP, dystonia is often underdiagnosed or misdiagnosed as spasticity, which can lead to a diagnostic delay of years.^4,5^ Therefore, there is a need for improving dystonia diagnosis in CP.

Despite an association with some common postures,^6^ the appearance of dystonia tends to vary based on the voluntary movement, body part involved, and the underlying etiology. This has necessitated an appropriately broad consensus definition which, unfortunately, cannot provide clear guidelines regarding the specific movement characteristics that may lead to a dystonia diagnosis in a given patient. This is in contrast to the consensus definition of spasticity which provides a clear exam maneuver (externally imposed stretch of a muscle at varying speed) that should yield an objective result (increased resistance at a certain joint angle or with increased speed of stretch).^7^ Validated dystonia diagnosis paradigms, like the Hypertonia Assessment Tool, provide clear exam maneuvers for eliciting dystonia but still require the practitioner to be adept at subjective assessment of any resultant involuntary movements or postures suggestive of dystonia, which can often be subtle.^2,8,9^ Given this, the gold standard dystonia diagnosis remains subjective expert identification, ideally in the form of consensus opinion of a panel of experts as is common clinical practice in large movement disorders centers. However, this form of diagnosis is not clinically feasible on a broad scale, particularly in centers where such specialists are not readily available.

Development of the current definition of dystonia and the HAT relied on expert consensus to identify key features of dystonia.^1,9^ However, they did not analyze discussions between experts as they were diagnosing dystonia in real-time. We hypothesize that distilling the key movement features that prompt experts to diagnose dystonia in a relatively homogenous CP patient population may be useful as a reference checklist for dystonia diagnosis in clinical practice. To this end, we conducted a qualitative thematic analysis of consensus-building discussions between three pediatric movement disorders physicians as they attempted to identify the presence or absence of dystonia in gait videos of 34 children with CP, a history of spasticity, and periventricular leukomalacia.

## Methods

This study was granted Human Subjects Research approval from the Washington University School of Medicine Institutional Review Board.

This was a retrospective case-control study of subjects who were seen in the St. Louis Children’s Hospital Cerebral Palsy Center from January 1, 2005 and December 31, 2018 (when the Center first started regularly collecting video during routine clinic visits). Our goal was to identify dystonia from videos taken of a relatively homogenous subject population engaging in the same functionally-relevant voluntary movement. This was done to make sure the engendered discussions could focus primarily on the aspects of movement most related to dystonia identification while controlling for other clinical factors. Standardization of the voluntary movement assessed was necessary as dystonia may appear differently if triggered by different voluntary movements. We also wanted to ensure that subjects had a history of spasticity, since co-morbid spasticity can make dystonia diagnosis particularly difficult in CP.^4^ Inclusion criteria were: 1) ICD-9 or ICD-10 diagnoses of spastic diplegic, spastic triplegic, spastic hemiplegic, or spastic quadriplegic CP, 2) Available video taken during routine clinic visits to the Cerebral Palsy Center, and 3) Presence of periventricular leukomalacia (PVL) on brain MRI per the clinical radiology report. Exclusion criteria were histories of either: 1) Traumatic brain injury (e.g. non-accidental trauma), 2) Brain surgeries, 3) Brain tumors, 4) Encephalitis, 5) Chemotherapy, 6) Metabolic disorders, and 7) Genetic disorders. Exclusion criteria were purposefully broad to focus on only a single type of brain injury, again to select as homogenous a patient population as possible. The videos of the remaining subjects were screened for clips of the subjects independently walking barefoot, with knees through toes visible, in a straight line towards the camera. This movement task was chosen for its functional relevance and because it allowed the best visualization of the whole subject during voluntary movement. Videos with subjects using hand-held walking aids were assessed, but videos with subjects walking in orthotics were not.

A single gait video clip from each included subject was de-identified and shared via a secure server with three fellowship-trained pediatric movement disorders physicians with particular interest in caring for children with CP (BRA, KU, and TP). All three completed pediatric neurology and movement disorders fellowship training at different institutions and all currently see patients in the St. Louis Children’s Hospital Cerebral Palsy Center. These putative CP motor phenotyping experts (henceforth referred to as “discussants”) watched the videos together in real-time and then silently voted regarding the presence or absence of dystonia in the lower extremities in each video. Discussants were asked to focus on identifying dystonia in the lower extremities as some subjects used hand-held walking aids for ambulation, precluding comparable assessment of upper extremity dystonia across all subjects. The results of the silent vote were then anonymously revealed and discussants then engaged in consensus-building discussion regarding how they approached identifying dystonia in the video. Discussants subsequently openly voted regarding their revised opinions on the presence or absence of dystonia in the video. Of note, given that the discussants all see patients in the Cerebral Palsy Center from which the videos used in this study were obtained, three of the subjects were recognized by at least one discussant. Codes generated from these videos were not included in further analysis.

Discussions were recorded, transcribed, and coded independently by two investigators (SS and LG) who were not present for the live discussion. Only codes shared between the two investigators were used for further analysis and for developing over-arching themes.

Comparisons of code frequency using Chi-square tests were made between four discussant categories based on initial and final votes regarding the presence of dystonia in each video: No to No, No to Yes, Yes to No, and Yes to Yes (Graph Pad Prism 8, GraphPad Software, San Diego, CA).

## Results

Seven hundred and twenty-one subjects carried ICD-9 or ICD-10 diagnosis of spastic CP, had video recorded of them during routine Cerebral Palsy Center clinic visits, and had at least one brain MRI clinical radiology report in their medical record documenting PVL. Of these subjects, 640 were excluded for having the disease processes or surgeries outlined above. The available videos of the remaining 81 subjects were screened with 40 subjects having videos of independent barefoot ambulation in a straight line towards the camera. A single gait video clip was selected for each of these 40 subjects and compiled together for further analysis. The subject clinical characteristics, video clip characteristics, and discussant decisions regarding presence or absence of dystonia in these clips are shown in Table 1. Subject age at the time of video recording ranged from 6 to 24 years old (average of 13.3 years old with 95% CI of 11.6-15.1 years old). Hand held walking aids were used in 25% of videos. Thirty percent of subjects (12/40) were categorized as Gross Motor Function Classification System^10^ (GMFCS) Level I, 40% (16/40) as GMFCS Level II, and 30% (12/40) as GMFCS Level III. No clinical data was missing from the included subjects.

**Table 1.** Subject characteristics

| Subject ID | Sex | Gestational age at birth (weeks) | Brain injury / CP etiology | GMFCS level | Video Details |  | Discussant Initial to Final Vote |  |  |
| --- | --- | --- | --- | --- | --- | --- | --- | --- | --- |
|  |  |  |  |  | Age (years) | Hand-held walking aid used | 1 | 2 | 3 |
| 1 | M | 30 | PVL | III | 16.4 | Y | Y to Y | Y to N | N to N |
| 2 | M | 25 | PVL | III | 18.8 | Y | N to N | N to N | N to N |
| 3 | F | Term | PVL (unknown etiology) | I | 6.7 | N | N to Y | Y to Y | N to Y |
| 4 | F | Premature (adopted) | PVL | I | 17.8 | N | N to N | N to N | N to N |
| 5 | M | 36 | HIE, PVL, IPH | III | 13.4 | N | N to Y | Y to Y | N to Y |
| 6 | F | Term | Stroke, PVL | II | 8.3 | N | Y to Y | N to Y | Y to Y |
| 7 | F | 25 | PVL | II | 12.8 | N | N to N | N to N | Y to N |
| 8 | F | 33 | PVL | III | 18.7 | Y | Y to Y | Y to Y | N to Y |
| 9 | M | 30 | PVL | III | 12.6 | Y | Y to Y | Y to Y | Y to Y |
| 10 | M | 36 | PVL (unknown etiology) | I | 8.3 | N | Y to Y | Y to Y | N to N |
| 11 | F | 27 | PVL | III | 17.7 | Y | N to Y | Y to Y | Y to Y |
| 12 | M | 30 | PVL | I | 3.6 | N | N to N | N to N | N to N |
| 13 | F | Premature (adopted) | Stroke, PVL | II | 18.7 | N | N to N | N to N | N to N |
| 14 | M | 28 | PVL | III | 19.1 | N | N to N | N to N | N to N |
| 15 | M | 31 | PVL | II | 15.5 | N | Y to Y | Y to Y | Y to Y |
| 16 | M | 31 | PVL | II | 16.1 | N | Y to Y | Y to Y | N to Y |
| 17 | F | 25 | PVL, IVH, PHH | II | 8.2 | N | Y to Y | Y to Y | Y to Y |
| 18 | M | 32 | PVL | II | 11.1 | N | Y to N | N to N | N to N |
| 19 | M | 27 | PVL | II | 11.1 | N | Y to N | Y to N | N to N |
| 20 | M | 33 | PVL | I | 9.5 | N | N to N | N to N | N to N |
| 21 | M | 35 | HIE, PVL | II | 8.9 | N | Y to Y | N to Y | Y to Y |
| 22 | F | Premature | PVL | II | 24.4 | N | N to N | N to N | Y to N |
| 23 | F | 24 | PVL | I | 22.3 | N | N to N | N to N | N to N |
| 24 | F | Premature (adopted) | PVL | I | 13.2 | N | Y to Y | N to N | N to N |
| 25 | F | 39 | HIE (placental abruption), PVL | II | 6.0 | N | Y to Y | Y to Y | Y to Y |
| 26 | F | 24 | PVL | II | 16.4 | N | N to N | N to N | Y to N |
| 27 | F | 26 | PVL, IVH | II | 4.4 | N | Y to Y | Y to Y | Y to Y |
| 28 | F | Term | PVL (unknown etiology) | III | 8.8 | Y | N to Y | Y to Y | Y to Y |
| 29 | M | 31 | PVL | III | 7.7 | Y | Y to N | N to N | N to N |
| 30 | F | Unknown (adopted) | PVL (unknown etiology) | I | 18.3 | N | N to N | N to Y | Y to Y |
| 31 | F | 28 | PVL | II | 6.2 | N | N to N | N to N | N to N |
| 32 | M | 27 | PVL, IVH | I | 9.2 | N | N to N | N to N | N to N |
| 33 | F | 28 | HIE (placental<br>abruption), PVL,<br>IVH | III | 12.9 | Y | Y to Y | Y to Y | Y to Y |
| 34 | F | 40 | PVL, IPH | I | 7.2 | N | N to Y | N to N | Y to Y |
| 35 | F | 28 | PVL, IVH | II | 23.2 | N | N to N | Y to N | N to N |
| 36 | F | 34 | HIE (placental<br>abruption), PVL | I | 17.6 | N | N to N | N to N | N to N |
| 37 | F | 40 | PVL (unknown<br>etiology) | II | 19.3 | N | N to N | N to N | N to N |
| 38 | M | 33 | PVL | I | 6.4 | N | Y to Y | Y to Y | N to Y |
| 39 | F | 28 | PVL | III | 13.9 | Y | N to N | N to N | N to N |
| 40 | M | 33 | PVL | III | 22.4 | Y | N to N | N to N | Y to Y |

HIE – hypoxic-ischemic encephalopathy, PVL – periventricular leukomalacia, IVH – intraventricular hemorrhage, IPH – intraparenchymal hemorrhage, PHH – post-hemorrhagic hydrocephalus, CP – cerebral palsy, GMFCS – Gross Motor Function Classification System<sup>10</sup>

Initial unanimous consensus regarding presence or absence of dystonia in the clips was only reached in 12/40 videos (30%). Following discussion, unanimous consensus was reached on an additional 22 videos (55%). Of these 34 videos, dystonia was unanimously identified in 15 (44%) and unanimously not identified in 19 (56%). No consensus was reached regarding the presence or absence of dystonia in 6 videos (15%). Discussants did not differ from each other in the frequency with which they initially or ultimately classified a video as having dystonia (Chi-square, p=0.992) (Figure 1).

**Figure 1.**
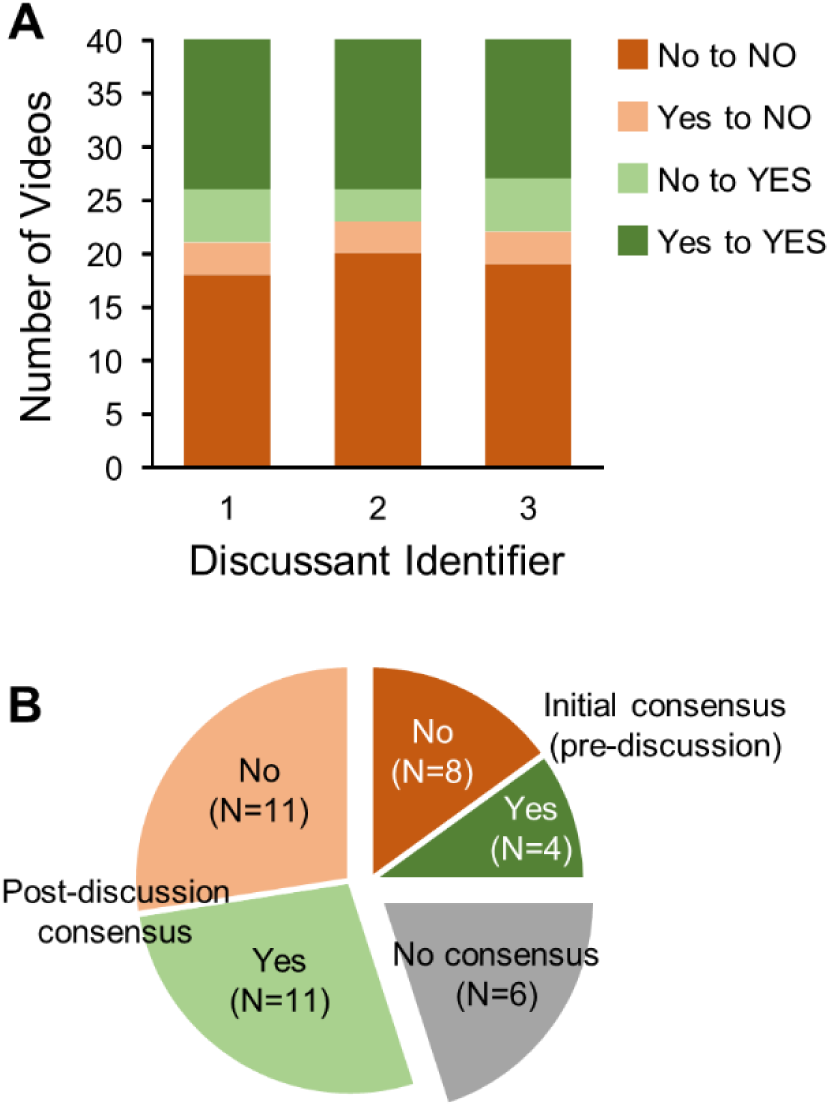
Voting results by discussant and across all videos. A) Frequencies of initial and final yes and no votes regarding the presence or absence of dystonia in subject videos, separated by discussant. B) Consensus voting results regarding the presence or absence of dystonia across all subject videos.

The consensus-building discussion transcript was ultimately parsed into 519 codes, representing 54 individual codes that were identified as occurring at least once. The most frequently used codes were expressions of uncertainty (e.g. “I think I see” or “I’m hedging”), movement variability over time (e.g. that movements happened only during a certain time period in the video but not at other times), and movement abnormalities that were identified as being unilateral, subtle (e.g. “a little bit” or “very mild”), in the foot, in the leg, or involving adduction (often explicitly noted but also inferred from statements like “crosses in” or “comes in”). The next most frequently used codes occurred after an apparent inflection point in the code frequency distribution and were also not as useful in defining movement qualities that could prompt dystonia identification. These codes included requests for additional chart information (e.g. “how old is she?” and “I want to know her diagnosis”), characterizing a movement as non-specifically abnormal (e.g. “something” or “funny”), and pondering whether the subject may have had a selective dorsal rhizotomy (Figure 2). Less frequent codes describing specific movements included foot inversion (N=6), foot plantar flexion (N=3), and leg extension (N=4).

**Figure 2.**
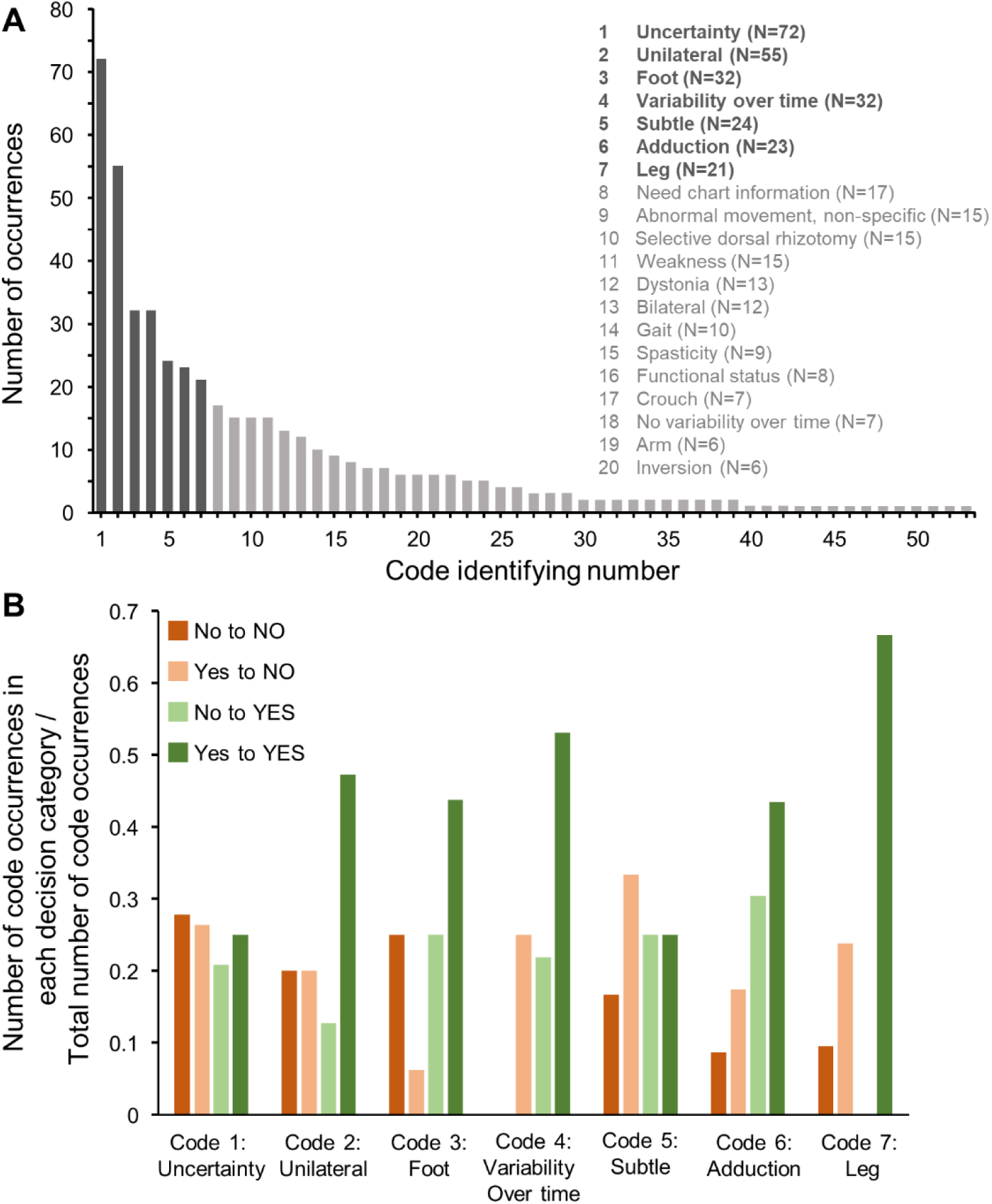
Coding of consensus-building discussions. A) Frequencies of individual codes (total of 54 individual codes with the top 20 codes indicated). There was a natural inflection point in the frequency distribution following the first 7 codes. B) Frequencies of top codes separated by discussant voting category. Codes were used at significantly different frequencies by discussants depending on how they voted regarding the presence or absence of dystonia in a video (Chi-square, p=0.004).

Codes were used at significantly different frequencies by discussants depending on how they voted regarding the presence or absence of dystonia in a video (Chi-square, p=0.004). Codes describing movements as being variable over time, unilateral, in the foot, in the leg, or involving adduction occurred more commonly when a discussant voted that dystonia was present in the video (Figure 2). These codes also commonly occurred together. At least three of these codes were present together in 18% of discussant’s statements containing any one of these codes, and all codes were present together in 7% of the statements. An example of a statement sharing all of these codes was: “I felt like her left foot intermittently turns in and that her right leg came in sharply once in the middle of the video.” Therefore, the primary theme derived from this study is that, when identifying dystonia, discussants cited: Unilateral foot or leg adduction that was variable over time. Codes attributable to this primary theme represented 31% of the total codes identified (163/519 codes). The secondary key theme was: Difficulty. Multiple codes documented the difficulty discussants had in identifying dystonia including: expressions of uncertainty, subtle movements, need for additional chart information, non-specific descriptors for abnormal movements, using the word “dystonia” or “dystonic” to define why dystonia was present, and the need to perform an exam. Codes attributable to this secondary theme also represented 31% of the total (159/519 codes).

## Discussion

Unilateral foot or leg adduction that is variable over time was the most commonly cited motor feature by fellowship-trained pediatric movement disorders physicians when identifying dystonia in gait videos of children and young adults with CP. Therefore, qualitative thematic analysis of motor phenotyping consensus-building discussions, as we have done in this study, could help identify specific motor features in homogenous patient populations that prompt dystonia identification. If done comprehensively, this could help objectively delineate specific movements to screen for when attempting to identify dystonia in children with CP.

Unilateral lower extremity abnormalities seemed to be more commonly cited when identifying dystonia despite the assessed voluntary movement (gait) involving both legs. This may be because the subjects were all affected by dystonia in one limb more than another (noting that their brain injuries were often asymmetric), because the limb engaging in the active swing portion of gait is more likely to demonstrate visualizable dystonia, or because difference in positioning between the limbs can best highlight the subtle movement abnormalities consistent with dystonia in these subjects.

Lower limb adduction was also frequently cited when identifying dystonia. Though sustained foot inversion is thought to be a dystonic posture, leg adduction has not been comparably described.^6^ In the context of the reviewed video clips, the leg adduction that may have prompted dystonia identification was likely variable over time and, therefore, dynamic and of short duration. If subtle and fleeting, leg adduction may have previously been missed as a movement critical for dystonia identification in children with CP. The concept of dystonic “scissoring” involving leg adduction has historically been noted in those with CP, but can be difficult to differentiate from spasticity.^11^ In these videos, it is possible that the variability in leg adduction over time allowed for differentiation from spasticity.

The difficulty of identifying dystonia in subjects with CP was also frequently cited, a phenomenon that has been described well in the literature.^4,5^ This difficulty is also highlighted by the fact that unanimous consensus was not reached regarding the presence or absence of dystonia in 15% of videos (6/40). However, when unanimous consensus was reached, dystonia was identified commonly (44%) in these subjects who were otherwise classified as having spastic CP. Therefore, though dystonia may not be the predominant form of tone in those with spastic CP, our results confirm that dystonia may still be commonly present and can emerge during a functionally important movement task like gait. Therefore, despite the difficulty in identifying it, it remains important to be vigilant for dystonia and its potential impact on function in all individuals with CP.

Our subjects were all independently ambulatory with or without an assistive device (GMFCS levels I-III^10^) and had diagnoses of spastic CP with PVL noted on their brain MRIs. Focusing on a relatively homogenous subject population likely aided extraction of discussion themes specific to dystonia identification. However, our results may only apply to this population. Conducting similar analyses with other groups of people with CP with different motor functional statuses, different patterns of brain injury, and different CP etiologies, may yield different movements that are most commonly associated with dystonia by motor phenotyping experts.

Choosing independent ambulation as the voluntary movement used to trigger dystonia was experimentally valuable for many reasons: 1) Gait is a functionally important voluntary movement, thus increasing the likelihood that any dystonia identified by motor phenotyping experts would have functional impact, 2) Gait is a relatively standardized movement across subjects that involves the whole body, allowing for the greatest likelihood of capturing dystonia in the video, and 3) Other standardized exam measures or voluntary movements were not reliably captured across all patients during routine clinic visits in this retrospective study. Future efforts could involve having motor phenotyping experts prospectively review video of standardized maneuvers (like performance of the HAT^2,8,9^) in subjects with CP. This might additionally alleviate some of the uncertainty expressed during consensus-building discussion regarding need for the exam to identify dystonia.

The motor phenotyping experts participating in this study are fellowship-trained pediatric movement disorders physicians who have trained in different locations but all currently practice in the same location. Though codes generated from videos where the subject was recognized by a discussant were excluded and though discussants were otherwise blinded to subject identity, there may still be some bias present when reviewing patient videos from your own practice center. Future studies could utilize online tele-consensus discussion between experts who practice at different centers.

Our results mark a critical first step in objectively delineating the specific movement features that are relevant to identifying dystonia in those with CP. Given that motor phenotyping has largely been seen as a subjective effort by putative experts, an analysis of the words used to describe movement disorders may contain our best hope of making what these experts do explicit. This will be beneficial both to teach others and for the education of the experts themselves regarding their own diagnostic practices. Furthermore, efforts to make dystonia diagnosis more objective are critical for ensuring this debilitating tone abnormality is diagnosed early and appropriately in those with CP.

## Data Availability

All data will be made available to qualified investigators upon request.

## Author Contributions

Dr. Aravamuthan was responsible for the design and conceptualization of the study, data analysis, data interpretation, and preparing the original draft of the manuscript. Dr. Ueda was responsible for the design and conceptualization of the study, data acquisition, and revising the manuscript for intellectual content. Mr. Miao, Ms. Smith, and Dr. Gilbert were responsible for data acquisition, analysis, and interpretation. Dr. Pearson was responsible for data acquisition and revising the manuscript for intellectual content.

